# Preventing aspiration events in hospital using prophylactic antiemetics: a systematic review and meta-analysis

**DOI:** 10.64898/2026.07.29.26357846

**Authors:** Kimberly B. Tworek, Marina Giovannoni, Janice Y. Kung, Vincent I. Lau

**Affiliations:** Department of Medicine, Faculty of Medicine and Dentistry, University of Alberta, Edmonton, Canada; University of Alberta, Geoffrey Robyn Sperber Health Sciences Library, Edmonton, Canada; Department of Critical Care Medicine, Faculty of Medicine and Dentistry, University of Alberta, Edmonton, Canada

## Abstract

**Introduction:** Aspiration events in hospitalized adults are common and linked to substantial morbidity, mortality, and healthcare burden. While antiemetics have been studied for aspiration prevention in specific settings, evidence for their broader preventive use in hospitalized adults is limited. We conducted a systematic review and meta-analysis to evaluate the effect of prophylactic scheduled antiemetics on aspiration events in adult inpatients.

**Methods:** Ovid MEDLINE, Ovid Embase, CINAHL, and Cochrane Library (via Wiley) were searched in May 2025. We included randomized controlled trials (RCTs) and observational studies assessing prophylactic scheduled antiemetic use in hospitalized adults, with aspiration events as the primary outcome. Secondary outcomes were aspiration pneumonia, hospital length of stay, increased oxygen requirement, ICU admission, and mortality. Two investigators screened and independently extracted data in duplicate using standardized data collection forms. Extracted information included study characteristics, patient demographics and clinical characteristics, interventions, and outcomes.

**Results:** Three RCTs met inclusion criteria (n = 515 patients; 237 antiemetic, 278 placebo). Pooled analysis of two RCTs demonstrated an 88% relative risk [RR] reduction of aspiration events; (2.8% [3/106] prophylactic antiemetic vs 27.9% [29/104] placebo; RR = 0.12, 95% confidence intervals [CI]: 0.02-0.73, p <0.0001, low certainty) in the antiemetic group. Pneumonia rates were also lower in the antiemetic group although they did not reach statistical significance (16.5% vs 32.4%; RR 0.44, 95% CI 0.15-1.28, P = 0.13, low certainty). ICU admission rates were lower in the antiemetic group (2.6% [2/76]) vs. placebo (23% [17/74]); RR = 0.11, 95% CI: 0.03-0.48, p=0.003, very low certainty). Mortality was similar between groups (35.0% vs 38.1%; RR 1.02, 95% CI: 0.84-1.24, P = 0.85, low certainty). No adverse events were reported among all studies.

**Conclusion:** Prophylactic scheduled antiemetics were associated with a significant reduction in aspiration and ICU admission. There was a non-significant but favorable trend towards reduction of pneumonia among hospitalized adults. Larger, high-quality trials are needed to clarify their role in aspiration prevention.

## INTRODUCTION

Aspiration of gastric contents or oropharyngeal secretions is a common and potentially serious complication in hospitalized adults, contributing to aspiration pneumonia, chemical pneumonitis, acute respiratory distress syndrome, and death (1,2), particularly in patients with impaired consciousness, dysphagia, neurological disorders or gastrointestinal dysfunction (3). Aspiration has been identified as an independent risk factor for increased hospital length of stay (4), along with increase in ICU admission, in-hospital mortality, and 30 day readmission rates (3–6), carrying a substantial financial burden and contributing to hospital capacity strain (4). Reported incidence rates vary widely depending on population and diagnostic criteria, but aspiration pneumonia alone accounts for a significant proportion of hospital-acquired infections. One large Japanese multicenter study found that aspiration pneumonia accounted for 38.4% of all pneumonia cases, with incidence increasing sharply with age and comorbidities (7).

Preventive strategies have traditionally emphasized nonpharmacologic measures, including head-of-bed elevation, dysphagia screening, and oral care protocols (8–10). However, pharmacologic interventions such as prophylactic scheduled antiemetic agents may have a role in reducing risk by decreasing nausea, vomiting, and subsequent aspiration events. The American Society of Anesthesiologists recommends antiemetics for patients at increased risk of postoperative nausea and vomiting, but not for routine aspiration prevention in healthy or low-risk patients (11). While antiemetics are routinely used in postoperative and chemotherapy settings, their potential to prevent aspiration across more general inpatient populations has not been systematically examined. Therefore, we conducted a systematic review and meta-analysis to assess whether prophylactic scheduled antiemetic use reduces aspiration events in this population.

## METHODS

### Protocol and Reporting Standards

This systematic review and meta-analysis was reported in accordance with the *Preferred Reporting Items for Systematic Reviews and Meta-Analyses (PRISMA) 2020* statement (12) and its protocol was prospectively registered in PROSPERO (CRD420251024973)(13).

### Eligibility Criteria

Inclusion criteria encompassed randomized controlled trials (RCTs) or observational studies that evaluated prophylactic scheduled antiemetics used to prevent aspiration events in adult patients (18 years or older) admitted to hospital. We excluded all experimental or animal studies and non–peer-reviewed reports. Non-research and conference abstracts, non-peer reviewed studies and websites were also excluded in addition to case reports and case series without control groups.

### Information Sources and Search Strategy

In collaboration with the research team, the medical librarian (JYK) developed and executed comprehensive searches in Ovid MEDLINE, Ovid Embase, CINAHL, and Cochrane Library (via Wiley) on May 20, 2025. To capture all relevant literature on preventing aspiration events in hospitals by using prophylactic antiemetics, relevant keywords and controlled vocabulary were carefully selected. There were no language or date limits applied. The full search strategy is provided in the Supplement.

### Screening and Full-Text Review Selection

All records were imported into Covidence (Veritas Health Innovations, Melbourne, Australia), and duplicates were excluded. Two reviewers (KBT and MG) independently screened titles and abstracts. We assessed relevance based on study design, population, and intervention characteristics. Articles that appeared potentially eligible were retrieved for full-text evaluation to confirm inclusion. Any discrepancies between reviewers were resolved through discussion, and when consensus could not be reached, a third reviewer (VL) provided adjudication. Reasons for study exclusion were documented during the full-text review phase.

### Data Extraction

Two reviewers (KBT and MG) independently extracted data in duplicate using standardized data collection forms. Any disagreements were resolved through discussion or, when necessary, by a third reviewer (VL) providing adjudication. Extracted information included study characteristics (design, setting), patient demographics, details of interventions (antiemetic class, timing, route, and dosing), and outcomes of interest (aspiration events, aspiration pneumonia, mortality, ICU admission). When essential data were missing, we contacted study authors to obtain the information.

### Outcomes

The primary outcome was witnessed or suspected aspiration events in hospital. Secondary outcomes included aspiration pneumonia, increased oxygen requirements and/or respiratory failure, acute respiratory distress syndrome (ARDS), hospital length of stay, transfer to ICU, ICU length of stay, and mortality. For each outcome, meta-analysis was only performed if there was sufficient data with at least 2 studies included.

### Risk of Bias Assessment

Two reviewers (KBT and MG) independently evaluated the risk of bias of included studies using the revised Cochrane risk-of-bias tool for randomized trials (RoB 2) (14), considering the following domains: randomization process, deviations from intended interventions, missing outcome data, measurement of outcome, selection of reported result. Each domain was rated as low, some concerns, or high risk of bias. The overall risk of bias for each study was determined by the highest risk rating across any individual domain.

### Effect Measures and Synthesis Methods

For dichotomous variables, risk ratios (RRs) and 95% confidence intervals (CIs). For continuous variables, mean differences (MDs) and 95% CIs were reported. Studies were pooled together using a random-effects model, weighted using the principle of inverse variance (15), where statistical heterogeneity was assessed using a random-effects model using the DerSimonian and Laird method, and 95% CIs was calculated by the Wald-type method using weighted mean differences. If no mean was reported, means were estimated from medians, ranges, rates, and sample sizes (16). All were calculated using RevMan Web 7.2.0 (The Cochrane Collaboration, London, United Kingdom).

### Certainty of Evidence

The overall certainty of the evidence was assessed using the Grading of Recommendations, Assessment, Development, and Evaluation (GRADE) framework (17). Following GRADE guidance, narrative summaries were used to convey effect sizes and certainty levels, with “probably” indicating moderate certainty, “may” indicating low certainty, and “uncertain” indicating very low certainty (18). Any discrepancies in risk of bias or GRADE assessments were resolved through discussion and consensus among the reviewers. For full GRADE evaluation, see Table 2.

## RESULTS

### Study Selection

The PRISMA flowchart is shown in Figure 1. Using the search strategy, a total of 3068 results were retrieved and after removing duplicates, 2720 unique results remained for the initial title and abstract screening. Of these, 3 randomized controlled trials met inclusion criteria. No observational studies met eligibility criteria.

**Figure 1:**
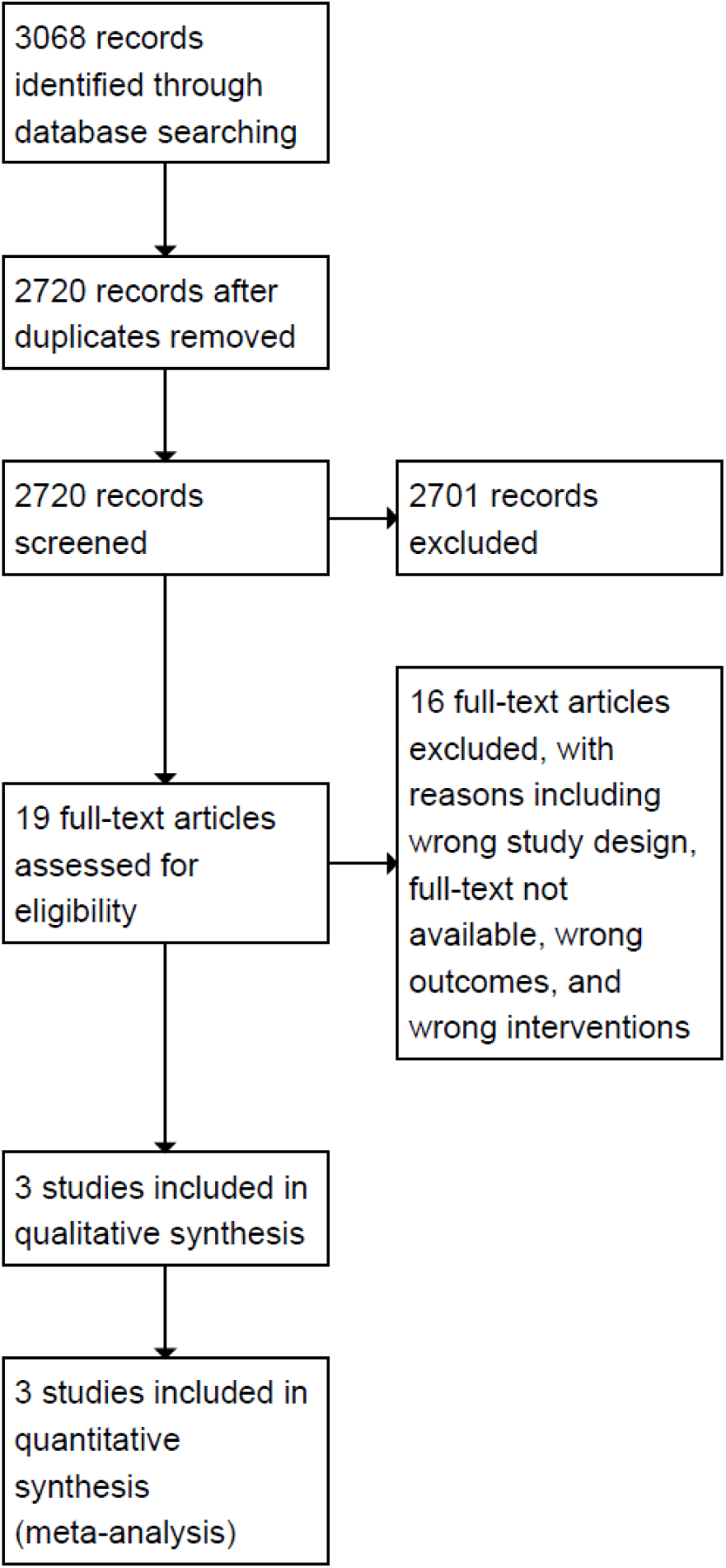
Flow Diagram.

### Study Characteristics

Three randomized controlled trials (RCTs) met inclusion criteria (19–21), with a total of 515 hospitalized adults, randomized to prophylactic antiemetics (n=237) versus placebo (n=278). The included trials were conducted in acute care hospital settings and evaluated prophylactic administration of antiemetic agents in adult patients admitted to hospital.

Detailed study characteristics are presented in Table 1. Dopamine antagonists (e.g. metoclopramide) were the antiemetic class chosen across all included studies given in standard dosing regimens. Control groups received placebo or standard care without routine antiemetic prophylaxis. Definitions of aspiration events were similar but not identical across studies and included witnessed aspiration, clinical suspicion based on acute respiratory compromise following emesis, or chest radiographic findings consistent with aspiration in the appropriate clinical context. Follow-up duration ranged from in-hospital monitoring to 21 days post-intervention.

**Table 1:** Study Characteristics.

| Study name<br>(year) country | Study design | Intervention<br>details | Population<br>(sample size:<br>intervention/cont<br>rol) | Outcomes with<br>available data | Outcome time<br>point | Method of<br>synthesis |
| --- | --- | --- | --- | --- | --- | --- |
| Allami (2022)<br>Iran | RCT | Domperidone 10<br>mg PO daily | Adults admitted<br>to hospital with<br>acute ischemic<br>stroke<br>(n = 150: 76/74) | Aspiration Events | Until hospital<br>discharge | MA |
|  |  |  |  | Aspiration<br>pneumonia<br>occurrence |  | MA |
|  |  |  |  | ICU Admission |  | Summary |
|  |  |  |  | Mortality |  | MA |
| Warusevitane<br>(2015)<br>UK | RCT | Metoclopramide<br>10 mg NG TID | Adults admitted<br>to hospital with<br>acute ischemic or<br>hemorrhagic<br>stroke with an NG<br>tube for >24<br>hours<br>(n = 60: 30/30) | Aspiration Events<br>Pneumonia<br>Mortality | 21 days | MA<br>MA<br>MA |
| Yavagal (2000)<br>India | RCT | Metoclopramide<br>10mg NG TID | Adults admitted<br>to the ICU with<br>an NG tube for<br>>24 hours<br>(n = 305:<br>131/174) | Pneumonia<br>Mortality | Until death or<br>discharge from<br>ICU | MA<br>MA |

**PO:** Per oral **NG:** Nasogastric **TID:** three times per day **MA:** meta-analysis of standardized effect sizes. **RCT:** randomized controlled trial.

### Risk of Bias in Studies

Overall risk of bias varied across studies. Two trials were judged to have low or probably low risk of bias across most domains, including adequate sequence generation, allocation concealment, and blinding of participants and outcome assessors (20, 21). One trial had some concerns due to unclear allocation concealment and incomplete outcome data (19). Selective outcome reporting was not clearly identified in any study. See supplement for ROB further detailed assessment.

### Results of Individual Studies and Syntheses

#### Primary Outcome: Aspiration Events

Two RCTs reported on aspiration events (n = 210; 106 antiemetic, 104 placebo) (19,21). Pooled analysis demonstrated an 88% relative risk [RR] reduction of aspiration events in those receiving prophylactic antiemetics (2.8%[3/106]) vs placebo (27.9% [29/104]); RR = 0.12, 95% confidence intervals [CI]: 0.02–0.73, p <0.0001, Risk Difference [RD] -25.1%, 95% CI -15.8 to - -34.5%, p<0.0001, low certainty) (Figure 2). Between-study heterogeneity was moderate (I² = 0.51), likely reflecting differences in patient populations, baseline aspiration risk, and antiemetic regimens.

**Figure 2:**
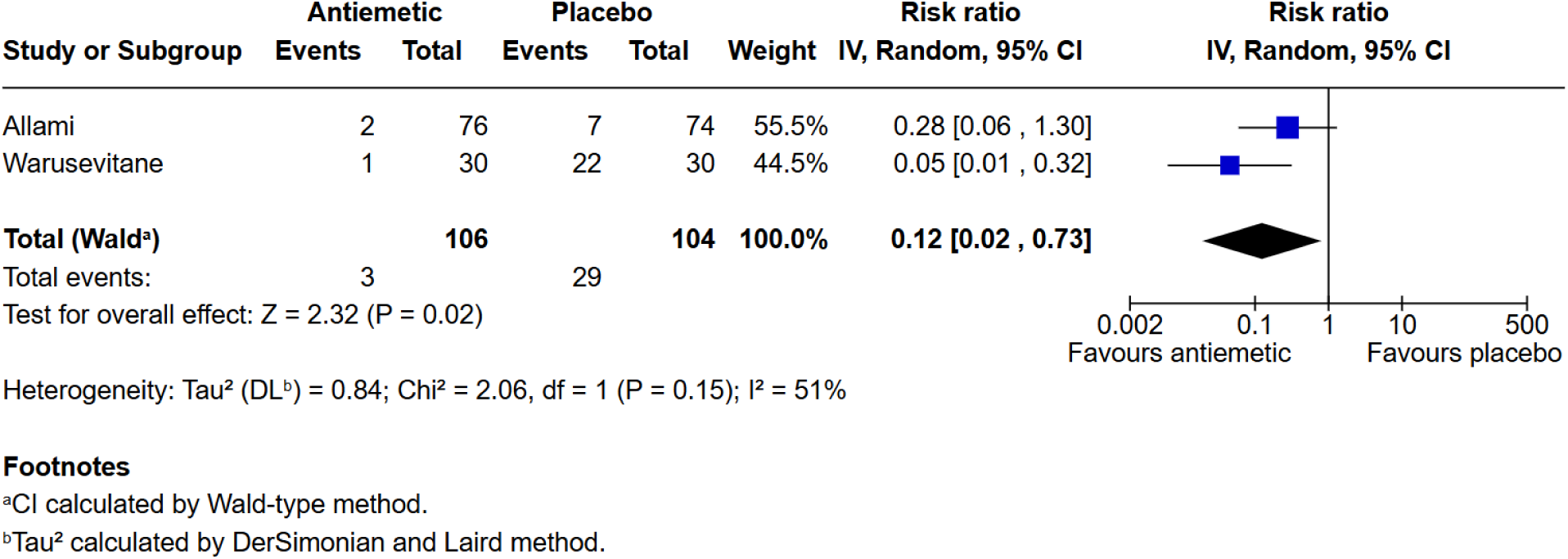
Aspiration Events.

### Secondary Outcomes

#### Pneumonia

Three RCTs reported rates of aspiration pneumonia (n = 509; 237 antiemetic, 278 placebo) (19–21). Though the reduction in pneumonia did not reach statistical significance, the antiemetic group showed a favorable trend compared with placebo (16.5% [39/237] vs 32.4% [90/278]); RR = 0.44, 95% CI: 0.15–1.28, p = 0.15; RD: -15.9%, 95% CI: -8.5% to -23.0%, p<0.0001, low certainty) (Figure 3).

**Figure 3:**
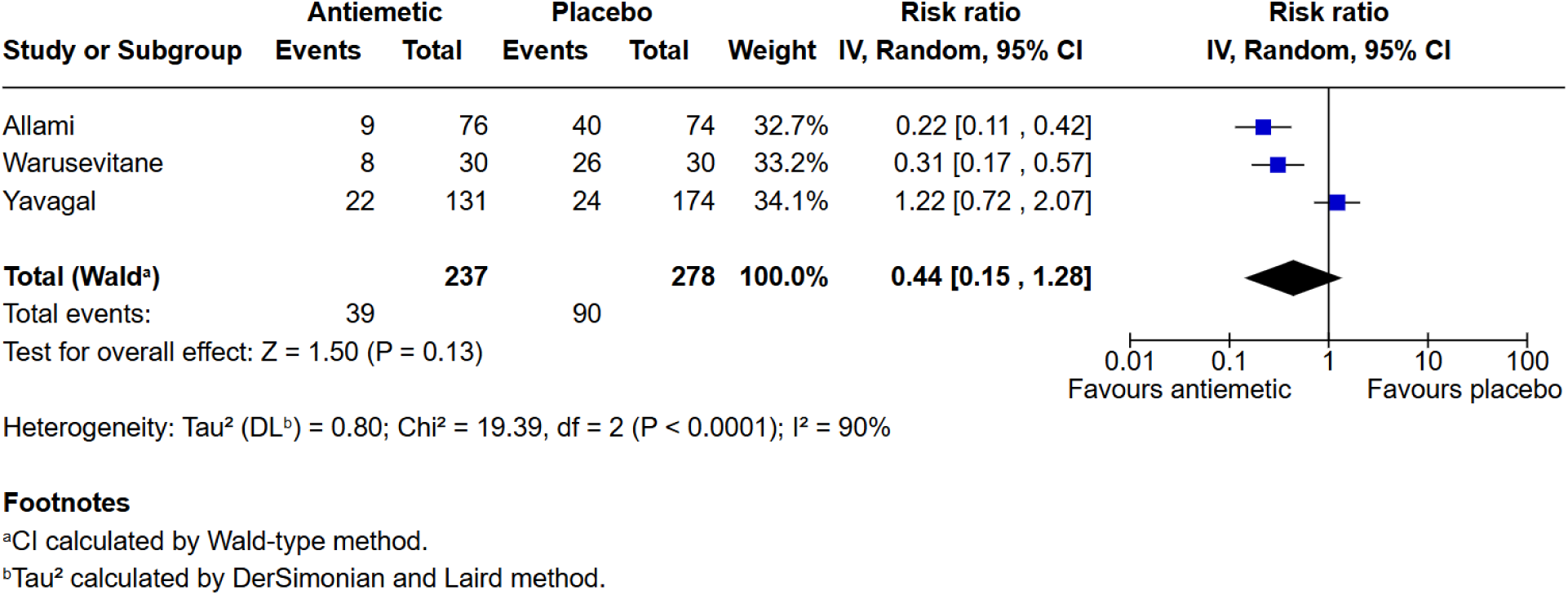
Pneumonia.

#### Mortality and Other Secondary Outcomes

All three trials reported all-cause mortality (n = 509; 237 antiemetic, 278 placebo). Mortality was similar between antiemetic (35.0% [83/237]) and placebo (38.1% [106/278] groups ; RR 1.02, 95% CI 0.84–1.24; RD: -3.1%, 95% CI -5.2% to 11.3%, p = 0.52, low certainty) (Figure 4). Unfortunately, data on ICU transfer, ICU length of stay, hospital length of stay, acute respiratory distress syndrome (ARDS), invasive ventilation, and escalation of oxygen requirements were inconsistently reported and could not be pooled due to heterogeneity in outcome definitions and incomplete reporting.

**Figure 4:**
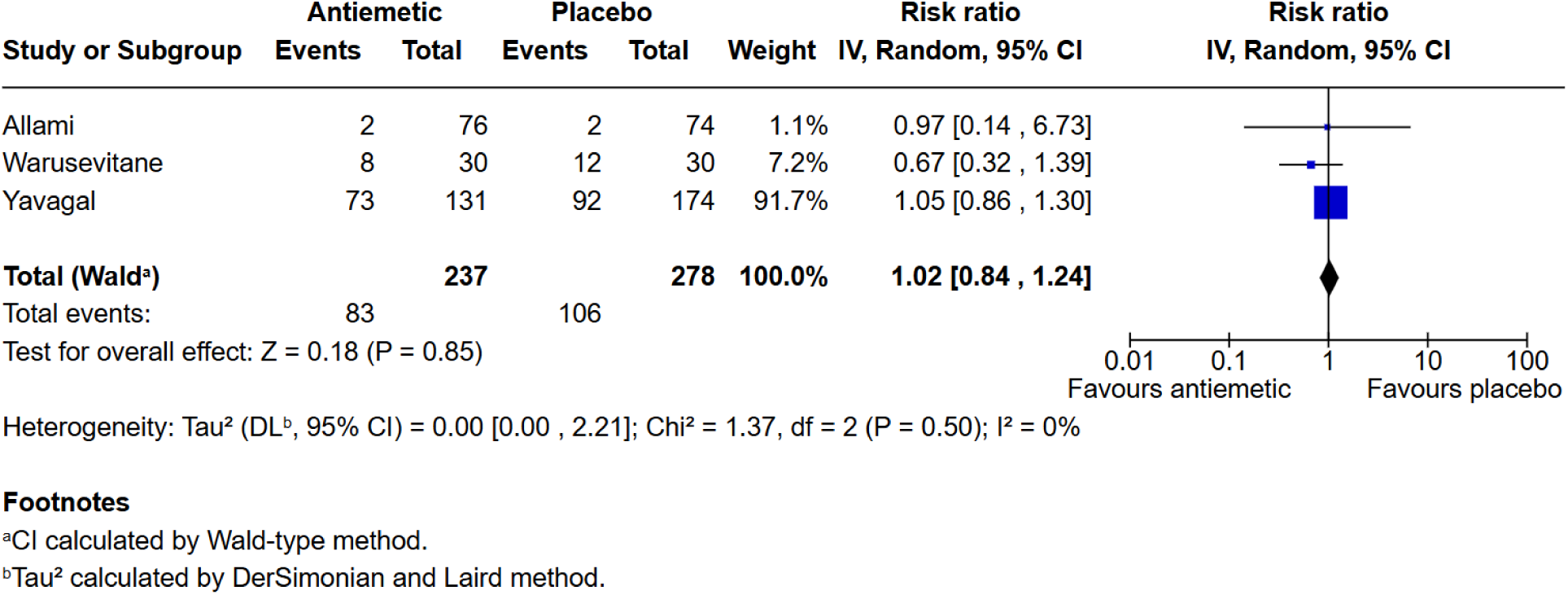
Mortality.

One study found ICU admission rates were significantly less in the antiemetic group (2.6% [2/76]) vs 23.0% [17/74] in the placebo group, RR 0.11, 95% CI 0.03 - 0.48, p = 0.003, RD: -20.3%, 95% CI: -10.0% to -31.3%, p<0.0001, low certainty) (21). Otherwise, individual studies did not demonstrate other statistically significant differences for any other outcomes mentioned between groups. See table 2 for summary of findings.

**Table 2:**
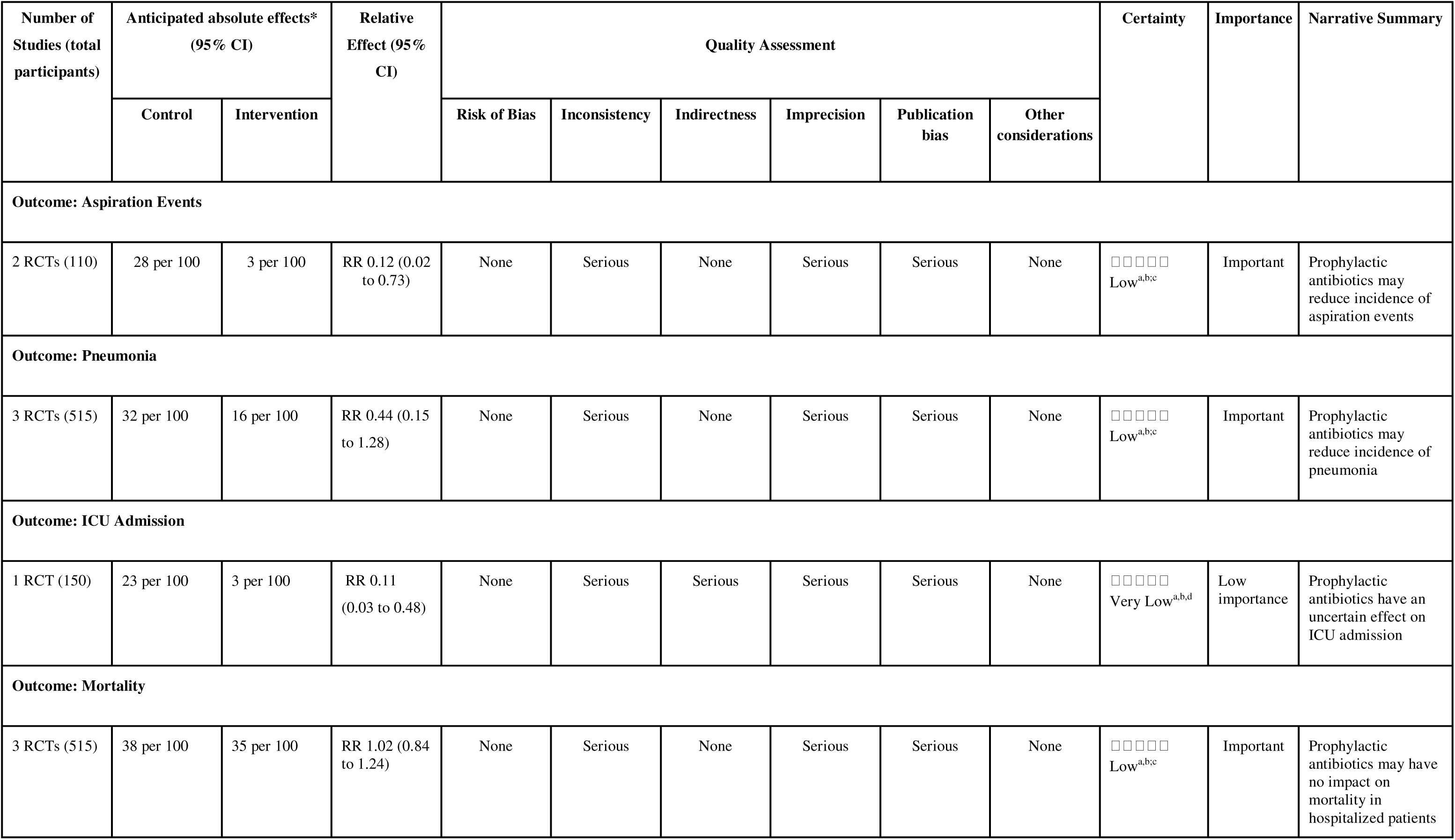
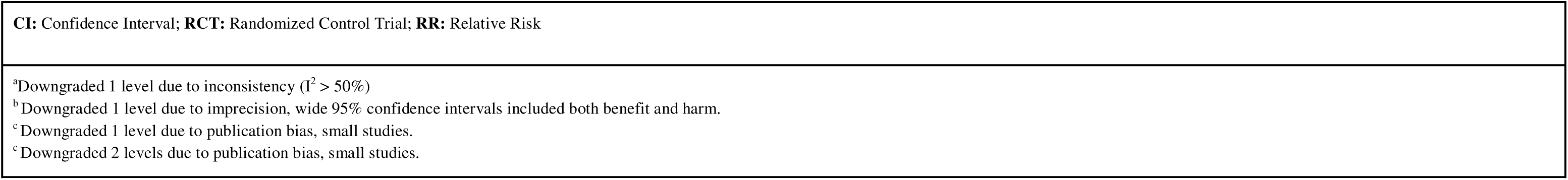
Summary of Findings with GRADE Assessment.

| Number of Studies (total participants) | Anticipated absolute effects* (95% CI) |  | Relative Effect (95% CI) | Quality Assessment |  |  |  |  |  | Certainty | Importance | Narrative Summary |
| --- | --- | --- | --- | --- | --- | --- | --- | --- | --- | --- | --- | --- |
|  | Control | Intervention |  | Risk of Bias | Inconsistency | Indirectness | Imprecision | Publication bias | Other considerations |  |  |  |
| Outcome: Aspiration Events |  |  |  |  |  |  |  |  |  |  |  |  |
| 2 RCTs (110) | 28 per 100 | 3 per 100 | RR 0.12 (0.02 to 0.73) | None | Serious | None | Serious | Serious | None | □□□□□<br>Low<sup>a,b,c</sup> | Important | Prophylactic antibiotics may reduce incidence of aspiration events |
| Outcome: Pneumonia |  |  |  |  |  |  |  |  |  |  |  |  |
| 3 RCTs (515) | 32 per 100 | 16 per 100 | RR 0.44 (0.15 to 1.28) | None | Serious | None | Serious | Serious | None | □□□□□<br>Low<sup>a,b,c</sup> | Important | Prophylactic antibiotics may reduce incidence of pneumonia |
| Outcome: ICU Admission |  |  |  |  |  |  |  |  |  |  |  |  |
| 1 RCT (150) | 23 per 100 | 3 per 100 | RR 0.11 (0.03 to 0.48) | None | Serious | Serious | Serious | Serious | None | □□□□□<br>Very Low<sup>a,b,d</sup> | Low importance | Prophylactic antibiotics have an uncertain effect on ICU admission |
| Outcome: Mortality |  |  |  |  |  |  |  |  |  |  |  |  |
| 3 RCTs (515) | 38 per 100 | 35 per 100 | RR 1.02 (0.84 to 1.24) | None | Serious | None | Serious | Serious | None | □□□□□<br>Low<sup>a,b,c</sup> | Important | Prophylactic antibiotics may have no impact on mortality in hospitalized patients |

|  |
| --- |
| **CI:** Confidence Interval; **RCT:** Randomized Control Trial; **RR:** Relative Risk |
| <sup>a</sup>Downgraded 1 level due to inconsistency ($I^2 > 50\%$)<br><sup>b</sup>Downgraded 1 level due to imprecision, wide 95% confidence intervals included both benefit and harm.<br><sup>c</sup>Downgraded 1 level due to publication bias, small studies.<br><sup>c</sup>Downgraded 2 levels due to publication bias, small studies. |

### Adverse Events

No study reported serious adverse events attributable to prophylactic antiemetic use (QTc prolongation, dystonia, constipation, etc.). Minor adverse effects, such as transient headache or constipation, were not systematically described.

### Certainty of Evidence

The certainty of evidence for aspiration events, pneumonia and mortality was judged to be low due to serious imprecision related to small sample size and limited event numbers, as well as concerns regarding potential publication bias. The evidence for ICU admission, derived from a single randomized controlled trial, was considered very low certainty due to serious imprecision and high risk of bias concerns inherent to single-study estimates. Overall, the certainty of evidence across outcomes was limited primarily by small sample sizes, heterogeneity in patient populations, and incomplete outcome reporting.

## DISCUSSION

In this systematic review and meta-analysis, prophylactic schedule antiemetic use was associated with a statistically significant 25% absolute risk and 88% relative risk reduction of aspiration events in hospital (19, 21). Although a similar directional effect was observed for aspiration pneumonia, this did not reach statistical significance, and mortality was unchanged. One study did describe a significant reduction in ICU admission among patients receiving prophylactic antiemetics compared with control, suggesting a potential reduction in clinical deterioration that requires higher levels of care. Overall, while limited by small sample size, these results could suggest potential benefit of prophylactic antiemetics for the prevention of aspiration events and their complications for hospitalized patients, warranting further investigation.

Current clinical guidelines focus primarily on nonpharmacologic preventive strategies such as head-of-bed elevation, dysphagia screening, and careful enteral feeding practices for the prevention of in-hospital aspiration events (2,8,22). The absence of strong evidence supporting routine prophylactic antiemetic use in general inpatient populations likely reflects the limited number and size of high quality trials. Our findings reaffirm this gap in knowledge and suggest that antiemetics may represent a complementary strategy to reduce aspiration risk and possibly prevent complication and/or progression to critical illness.

Reducing aspiration events is clinically important given their substantial impact on both patient outcomes and healthcare systems. Aspiration can lead to respiratory compromise, increased oxygen requirements, additional diagnostic investigations, prolonged hospitalization, ICU admission, mechanical ventilation, and increased healthcare costs (2, 4, 6). Furthermore, aspiration events often affect patient disposition and discharge planning, as ongoing aspiration risk may delay transfer to lower-acuity settings or discharge home. These consequences are particularly relevant among medically complex patients with impaired airway protection, altered consciousness, neurological disease, or enteral feeding requirements. In critically ill and mechanically ventilated patients, where aspiration is exceedingly common (23,24), even modest reductions in aspiration events could translate into meaningful reductions in morbidity and resources. The observed reduction in ICU admission in one included study may reflect prevention of aspiration-related respiratory deterioration. This is biologically plausible given antiemetics aim to reduce nausea, vomiting, and regurgitation, thereby theoretically decreasing airway exposure to gastric contents. However, implementation of routine prophylactic antiemetic therapy in hospitalized patients would also require careful consideration of safety, monitoring burden, and unintended consequences. Although no included study reported significant adverse events such as QTc prolongation or associated arrhythmias including Torsades de Pointes, these findings should be interpreted cautiously given the limited sample size and inconsistent adverse event reporting. Antiemetic agents, particularly dopamine antagonists and 5-HT3 antagonists, are associated with known risks including QT prolongation, extrapyramidal symptoms, delirium, constipation, and electrolyte abnormalities (25). Widespread implementation of prophylactic antiemetics could therefore increase requirements for ECG monitoring, electrolyte surveillance, and electrolyte replacement protocols, particularly among medically complex inpatients already at elevated arrhythmogenic risk. Importantly, the potential benefit of aspiration prevention must be balanced against the possibility of introducing other clinically significant complications, including cardiac arrhythmias and cardiac arrest. Reassuringly, this systematic review and meta analysis, there was no increased mortality or adverse events observed from antiemetic use which may demonstrate a signal that patients did not experience these clinically significant complications, however the overall risk-benefit profile of prophylactic antiemetic therapy will likely vary depending on baseline aspiration risk, comorbidity burden, and antiemetic agent selection.

This study has several strengths. We performed a comprehensive search based on PRSIMA guidelines across multiple databases without language or date restrictions, applied duplicate screening and data extraction, and assessed risk of bias and certainty of evidence using standardized tools (14, 18). However, important limitations should be considered. First, only three RCTs met inclusion criteria, resulting in a very low sample size and limiting the evidence base and of these three all used anti-dopaminergic medications, which may not be the typical antiemetic medication used in hospital (i.e anti-serotonergic medications such as ondansetron). Second, heterogeneity in patient populations, intervention protocols, and outcome definitions may have influenced pooled estimates, in addition to not all outcomes of interest being universally reported. Patient populations were also not generalizable for entire hospitalization populations, with 2 of 3 studies reporting on patients who had suffered strokes. Finally, aspiration events and resultant pneumonia are challenging to diagnose reliably given no standardized measure making misclassification possible.

Future research should prioritize large, adequately powered multicenter RCTs targeting clearly defined high-risk inpatient populations, such as those with impaired consciousness, advanced neurological disease, or enteral feeding. Standardized definitions of aspiration events and aspiration pneumonia, along with systematic adverse event reporting, will be essential to gaining further clarity of the role of prophylactic antiemetics in aspiration prevention.

## CONCLUSION

In hospitalized adults, prophylactic antiemetic use was associated with a significant reduction in aspiration events, low certainty of evidence. While limited by small sample size, a favorable trend was observed in the reduction of aspiration pneumonia. Prophylactic antiemetics may offer benefit in selected high-risk populations, however current evidence is insufficient to support routine prophylactic use for aspiration prevention in general inpatient settings. Larger, high-quality RCTs are needed to clarify their role in aspiration prevention.

## Data Availability

All data produced in the present study are available upon reasonable request to the authors

