## Supplemental Materials for "Preventing aspiration events in hospital using prophylactic antiemetics: a systematic review and meta-analysis"

Table 1s: PRISMA Checklist

| **Section and Topic** | **Item #** | **Checklist item** | **Location where item is reported** |
| --- | --- | --- | --- |
| **TITLE** | | |  |
| Title | 1 | Identify the report as a systematic review. | 1, 4 |
| **ABSTRACT** | | |  |
| Abstract | 2 | See the PRISMA 2020 for Abstracts checklist. | 2 |
| **INTRODUCTION** | | |  |
| Rationale | 3 | Describe the rationale for the review in the context of existing knowledge. | 4 |
| Objectives | 4 | Provide an explicit statement of the objective(s) or question(s) the review addresses. | 4 |
| **METHODS** | | |  |
| Eligibility criteria | 5 | Specify the inclusion and exclusion criteria for the review and how studies were grouped for the syntheses. | 5 |
| Information sources | 6 | Specify all databases, registers, websites, organisations, reference lists and other sources searched or consulted to identify studies. Specify the date when each source was last searched or consulted. | 5 |
| Search strategy | 7 | Present the full search strategies for all databases, registers and websites, including any filters and limits used. | 5, Supplement for full search strategy |
| Selection process | 8 | Specify the methods used to decide whether a study met the inclusion criteria of the review, including how many reviewers screened each record and each report retrieved, whether they worked independently, and if applicable, details of automation tools used in the process. | 5,6 |
| Data collection process | 9 | Specify the methods used to collect data from reports, including how many reviewers collected data from each report, whether they worked independently, any processes for obtaining or confirming data from study investigators, and if applicable, details of automation tools used in the process. | 6 |
| Data items | 10a | List and define all outcomes for which data were sought. Specify whether all results that were compatible with each outcome domain in each study were sought (e.g. for all measures, time points, analyses), and if not, the methods used to decide which results to collect. | 6 |
|  | 10b | List and define all other variables for which data were sought (e.g. participant and intervention characteristics, funding sources). Describe any assumptions made about any missing or unclear information. | 6 |
| Study risk of bias assessment | 11 | Specify the methods used to assess risk of bias in the included studies, including details of the tool(s) used, how many reviewers assessed each study and whether they worked independently, and if applicable, details of automation tools used in the process. | 6 |
| Effect measures | 12 | Specify for each outcome the effect measure(s) (e.g. risk ratio, mean difference) used in the synthesis or presentation of results. | 7 |
| Synthesis methods | 13a | Describe the processes used to decide which studies were eligible for each synthesis (e.g. tabulating the study intervention characteristics and comparing against the planned groups for each synthesis (item #5)). | 7 |
|  | 13b | Describe any methods required to prepare the data for presentation or synthesis, such as handling of missing summary statistics, or data conversions. | 6,7 |
|  | 13c | Describe any methods used to tabulate or visually display results of individual studies and syntheses. | 7 |
|  | 13d | Describe any methods used to synthesize results and provide a rationale for the choice(s). If meta-analysis was performed, describe the model(s), method(s) to identify the presence and extent of statistical heterogeneity, and software package(s) used. | 7 |
|  | 13e | Describe any methods used to explore possible causes of heterogeneity among study results (e.g. subgroup analysis, meta-regression). | 7 |
|  | 13f | Describe any sensitivity analyses conducted to assess robustness of the synthesized results. |  |
| Reporting bias assessment | 14 | Describe any methods used to assess risk of bias due to missing results in a synthesis (arising from reporting biases). | 7 |
| Certainty assessment | 15 | Describe any methods used to assess certainty (or confidence) in the body of evidence for an outcome. | 7 |
| **RESULTS** | | |  |
| Study selection | 16a | Describe the results of the search and selection process, from the number of records identified in the search to the number of studies included in the review, ideally using a flow diagram. | 7,8 |
|  | 16b | Cite studies that might appear to meet the inclusion criteria, but which were excluded, and explain why they were excluded. | 7, 8, Fig 1 |
| Study characteristics | 17 | Cite each included study and present its characteristics. | 8, Table 1 |
| Risk of bias in studies | 18 | Present assessments of risk of bias for each included study. | 8, Supplement |
| Results of individual studies | 19 | For all outcomes, present, for each study: (a) summary statistics for each group (where appropriate) and (b) an effect estimate and its precision (e.g. confidence/credible interval), ideally using structured tables or plots. | 8, 9, Table 2 |
| Results of syntheses | 20a | For each synthesis, briefly summarise the characteristics and risk of bias among contributing studies. | 8, 9, Table 2 |
|  | 20b | Present results of all statistical syntheses conducted. If meta-analysis was done, present for each the summary estimate and its precision (e.g. confidence/credible interval) and measures of statistical heterogeneity. If comparing groups, describe the direction of the effect. | 8, 9, Table 2 |
|  | 20c | Present results of all investigations of possible causes of heterogeneity among study results. |  |
|  | 20d | Present results of all sensitivity analyses conducted to assess the robustness of the synthesized results. |  |
| Reporting biases | 21 | Present assessments of risk of bias due to missing results (arising from reporting biases) for each synthesis assessed. | 8, 9, Table 2 |
| Certainty of evidence | 22 | Present assessments of certainty (or confidence) in the body of evidence for each outcome assessed. | 10, Table 2 |
| **DISCUSSION** | | |  |
| Discussion | 23a | Provide a general interpretation of the results in the context of other evidence. | 10, 11 |
|  | 23b | Discuss any limitations of the evidence included in the review. | 13 |
|  | 23c | Discuss any limitations of the review processes used. | 13 |
|  | 23d | Discuss implications of the results for practice, policy, and future research. | 12, 13 |
| **OTHER INFORMATION** | | |  |
| Registration and protocol | 24a | Provide registration information for the review, including register name and registration number, or state that the review was not registered. | 5 |
|  | 24b | Indicate where the review protocol can be accessed, or state that a protocol was not prepared. | 5 |
|  | 24c | Describe and explain any amendments to information provided at registration or in the protocol. |  |
| Support | 25 | Describe sources of financial or non-financial support for the review, and the role of the funders or sponsors in the review. | 1 |
| Competing interests | 26 | Declare any competing interests of review authors. | 1 |
| Availability of data, code and other materials | 27 | Report which of the following are publicly available and where they can be found: template data collection forms; data extracted from included studies; data used for all analyses; analytic code; any other materials used in the review. | Supplement |

*From:* Page MJ, McKenzie JE, Bossuyt PM, Boutron I, Hoffmann TC, Mulrow CD, et al. The PRISMA 2020 statement: an updated guideline for reporting systematic reviews. BMJ 2021;372:n71. doi: 10.1136/bmj.n71. This work is licensed under CC BY 4.0. To view a copy of this license, visit <https://creativecommons.org/licenses/by/4.0/>

Table 2s: Search Strategies

| **Database** | **Search Strategy** |
| --- | --- |
| **MEDLINE**  Ovid MEDLINE(R) ALL 1946 to May 19, 2025 | 1. exp Deglutition Disorders/ or Deglutition/  2. exp Pneumonia, Aspiration/  3. exp Respiratory Aspiration/  4. (aspiration pneumon* or pneumonit* or pulmonary aspiration* or foreign body aspiration or aspiration event*).mp.  5. ((airway* or respirat*) adj3 aspirat*).mp.  6. ((swallow* or deglutit* or dysphag*) adj3 (abnormal* or condition* or damage* or disturbance* or disorder* or difficult* or dysfunction* or impair* or injur*)).tw,kf.  7. ((throat or oesophag* or esophag* or pharyn* or oropharyn*) adj3 (abnormal* or condition* or damage* or disturbance* or disorder* or difficult* or dysfunction* or impair* or injur*)).tw.  8. or/1-7  9. exp Antiemetics/ or (antiemetic* or anti-emetic*).mp.  10. exp Dopamine Antagonists/  11. (dopamin* adj2 antagonist*).tw,kf.  12. (chlorpromazine or aminazine or chloractil or chlordelazine or contomin or dozine or fenactil or largactil or ormazine or propaphenin or thorazine).mp.  13. (domperidon* or domidon or evoxin or gastrocure or motilium or motillium or motinorm or costi or nauzelin).mp.  14. (metoclopramide or cerucal or clopra or degan or gastrese or gastrobid continus or gastroflux or gastromax or maxolon or maxeran or metaclopramide or metozolv or metramid or migravess or mygdalon or octamide or parmid or primperan or pylomid or reglan or reliveran or rimetin).mp.  15. (haloperidol or dozic or Aloperidin or Bioperidolo or Brotopon Duraperidol or fortunan or haldol or kentace or Einalon or Eukystol, or Halosten or Keselan or Linton or Peluces or Serenase or Sigaperidol or serenace).mp.  16. (prochlorperazine or buccastem or compazine or compro or emezine or procot or proziere or Phenotil or stemetil or Stemzine).mp.  17. (promethazine or Avomine or adgan or aler-dryl or aler-tab or aller-dryl topical or allergia or allermax or altaryl or anergan or antihist or antinaus or antituss or atosil or banaril or banophen or beldin or belix or ben tann or benadryl or benahist or bendylate or benekraft or benzhydramine or bromanate or calm-aid or derma-pax or dimedrol or dimine or diphen or diphenadryl or diphenhist or diphenhydramine or diphenmax or diphenyl or diphergan or diprazin or dormarex or dytan or dytuss or eldadryl or Fargan or Farganesse or genahist or hydramine or hyrexin or isopromethazine or Lergigan or medinex or nervine or nightcalm or nu-med or nytol or pardryl or paxidorm or pediacare or pentazine or phenadoz or phenazine or phendry or phenergan or phenerzine or phenoject or phensedyl or phenylbenzene or pipolphen or pro-med or proazamine or progan or promacot or promet or prometazin or promethegan or prorex or prothazin or Prothiazine or provigan or pyrethia or quenalin or remsed or Romergan or Receptozine or rumergan or siladryl or siladyl or silphen or sleep tab* or sleep-ettes or sleep-eze or sleepia or sleepinal or sominex or somnicaps or trux-adryl or tusstat or twilite or uni-hist or uni-tann or unisom sleepgels or unisom sleepmelts or valu-dryl or wehdryl or zipan).mp.  18. exp Serotonin Antagonists/  19. (serotonin adj2 antagonist*).tw,kf.  20. (dolasetron or anzemet).mp.  21. (granisetron or granisol or kytril or sancuso).mp.  22. (Ondansetron or zensana or zofran).mp.  23. (tropisetron or Navoban or Setrovel).mp.  24. exp Cholinergic Antagonists/ or anticholinergic agent*.tw.  25. (scopolamine or atrochin or boroscopol or buscapine or buscolysin or buscopan or butylscopolamine or butylscopolammonium bromide or epoxytropine tropate or hyocine hydrobromide or hyoscinbutylbromide or hyoscine or kwell or levo-duboisine or maldemar or scoburen or scopace or scopoderm or scopolaminebutylbromide or scopolaminum hydrobromicum or scopolan or transderm or transderm-scop or travacalm or vorigeno).mp.  26. exp Histamine Antagonists/ or (histamine antagonist* or antihistamine* or anti-histamine*).mp.  27. (buclizine or cyclizine).tw.  28. (dimenhydrinate or antimo or aviomarin or biodramina or cinfamar or contramareo or dimen heumann or dimen lichtenstein or dimetabs or dinate or diphenhydramine theoclate or dramamine or dramin or Driminate or dramanate or dramoject or dymenate or gravol or Gravamin or marmine or nausicalm or reisegold or reisetabletten ratiopharm or reisetabletten stada or rodovan or rubiemen or superpep or travel-eze or travel-wise or triptone or uni-calm or Vertirosan or Viabom or vomex or vomacur or vomisin or wehamine).mp.  29. (Trimethobenzamide or barogan or benzacot or tebamide or ticon or tigan).mp.  30. (meclizine or agyrax or antivert or bonamine or bonikraft or bonine or chiclida or histametizyn or meclicot or meclozine or medivert or parachloramine or ruvertm or univert).mp.  31. exp Benzodiazepines/ or benzodiazepine*.mp.  32. (lorazepam or almazine or apolorazepam or ativan or donix or durazolam or idalprem or laubeel or lorazep or novolorazem or nuloraz or orfidal wyeth or sedicepan or sinestron or somagerol or tolid or temesta).mp.  33. exp Adrenal Cortex Hormones/  34. corticosteroid*.tw,kf.  35. (dexamethasone or aacidexam or adexone or adrenocot or aeroseb or aknichthol dexa or alba-dex or alin or ambene or amplidermis or anemul mono or antimicotico or aquapred or auricularum or auxiloson or azona or baycadron or baycuten or cebedex or corson or cortastat or cortidex or cortidexason or cortisumman or corto-tavegil or dalalone or deca or decacort or decaderm or decadron or decalix or decasone or decaspray or dectancyl or deenar or dekasol or deronil or desamethasone or desameton or dexa-mamallet or dexa-rhinosan or dexa-scheroson or dexa-sine or dexacen or dexacort phosphate or dexacort* or dexafarma or dexafluorene or dexair or dexaject or dexalocal or dexamecortin or dexameth or dexamethasonedisodium phosphate or dexamethasonum or dexamonozon or dexapos or dexasol or dexasone or dexinoral or dexium or dexpak or dinormon or doxiproct or fluorodelta or fortecortin or gammacorten or hexadecadrol or hexadrol or lokalison or loverine or maxidex or medidex or metazone or methylfluorprednisolone or millicorten or mymethasone or ocasa or ocu-dex or oradexon or orgadrone or otomize or ozurdex or predni or primethasone or robadex or soludecadron or solurex or spersadex or trabit or visumetazone or voren).mp.  36. (methylprednisolone or a-methapred or adlone or ak-pred or ak-tate or aprednislon or articulose or asmacortone or balpred or blephamide liquifilm or bubbli-pred or caberdelta or capsoid or codelson or cortalone or corti-clyss or cortimed or cortisolone or cotolone or cryosolona or decaprednil or decortin or delta-cortef or delta-diona or delta-phoricol or deltacortilen or deltacortril or deltahydrocortisone or deltasolone or deltastab or deltidrosol or depmedalone or depo moderin or depo-medrol or depo-nisolone or depoject or depopred or dhasolone or di-adreson-f or diopred or dontisolon or duralone or duro cort or econopred or emmetipi or esametone or estilsona or firmacort or fisopred or flo-pred or frisolona or gupisone or hexacortone or hostacortin or hydeltra or hydeltrasol or hydrocortancyl or inf-oph or inflamase or inflanefran or isolone or key-pred or klismacort or kuhlprednon or lenisolone or lepi-cortinolo or locaseptil or longiprednil or med-jec or medicort or medlone or medrate or medrol or medrone or mega-star or meprdl or meprolone or metacortandralone or methacort or methylcotol or methylcotolone or methylone or methylpred or methylprednisolonum or meti derm or meticortelone or metilbetasone solubile or metipred or metrocort or metypresol or metysolon or millipred or ocu-pred or omnipred or ophtho-tate or opredsone or orapred or panafcortelone or pediapred or poly-pred liquifilm or polypred or precortalon aquosum or precortisyl or pred-clysma or pred-ject or pred-phosphate or predacorten or predair or predaject or predalone or predate or predcor or predeltilone or predenema or predfoam or predicort or predmix or prednabene or prednefrin or predni-coelin or predni or predni-helvacort or predni-m-tablinen or predni-pos or prednicortelone or prednihexal or prednilen or predniocil or prednisol or prednisolone or prednoral or predonine or predsol or prelone or pri-cortin or pri-methylate or pricortin or radilem or sano-drol or scherisolone-kristall or sieropresol or solpredone or solu moderin or solu-medrol or sterane or stintisone or summicort or urbason or urbasonsoluble or veripred or wyacort).mp.  37. exp cannabinoids/ or cannabinoid*.tw,kf.  38. (cannador or charas or ganja* or hashish or hemp or cannabis or marihuana or marijuana).tw,kf.  39. (marinol or dronabinol or tetrahydrocannabinol).mp.  40. or/9-39  41. (prevent* or mitigat* or minimi?* or avoid*).tw.  42. prophyla*.tw,kf.  43. 41 or 42  44. 8 and 40 and 43  45. animals/ not (animals/ and humans/)  46. (veterinary or rabbit or rabbits or animal or animals or mouse or mice or rodent or rodents or rat or rats or murine or hamster* or pig or pigs or piglets or swine or porcine or horse* or equine or cow or cows or cattle or bovine or goat or goats or sheep or lambs or ovine or monkey or monkeys or trout or marmoset$1 or canine or dog or dogs or feline or cat or cats or zebrafish).ti.  47. 45 or 46  48. 44 not 47 |
| **Embase**  Ovid Embase 1974 to 2025 May 19 | 1. exp aspiration pneumonia/  2. exp acid aspiration/  3. (aspiration pneumon* or pneumonit* or pulmonary aspiration* or foreign body aspiration or aspiration event*).mp.  4. ((airway* or respirat*) adj3 aspirat*).mp.  5. ((swallow* or deglutit* or dysphag*) adj3 (abnormal* or condition* or damage* or disturbance* or disorder* or difficult* or dysfunction* or impair* or injur*)).tw,kw.  6. ((throat or oesophag* or esophag* or pharyn* or oropharyn*) adj3 (abnormal* or condition* or damage* or disturbance* or disorder* or difficult* or dysfunction* or impair* or injur*)).tw.  7. or/1-6  8. exp antiemetic agent/ or (antiemetic* or anti-emetic*).mp.  9. exp dopamine receptor blocking agent/  10. (dopamin* adj2 antagonist*).tw,kw.  11. (chlorpromazine or aminazine or chloractil or chlordelazine or contomin or dozine or fenactil or largactil or ormazine or propaphenin or thorazine).mp.  12. (domperidon* or domidon or evoxin or gastrocure or motilium or motillium or motinorm or costi or nauzelin).mp.  13. (metoclopramide or cerucal or clopra or degan or gastrese or gastrobid continus or gastroflux or gastromax or maxolon or maxeran or metaclopramide or metozolv or metramid or migravess or mygdalon or octamide or parmid or primperan or pylomid or reglan or reliveran or rimetin).mp.  14. (haloperidol or dozic or Aloperidin or Bioperidolo or Brotopon Duraperidol or fortunan or haldol or kentace or Einalon or Eukystol, or Halosten or Keselan or Linton or Peluces or Serenase or Sigaperidol or serenace).mp.  15. (prochlorperazine or buccastem or compazine or compro or emezine or procot or proziere or Phenotil or stemetil or Stemzine).mp.  16. (promethazine or Avomine or adgan or aler-dryl or aler-tab or aller-dryl topical or allergia or allermax or altaryl or anergan or antihist or antinaus or antituss or atosil or banaril or banophen or beldin or belix or ben tann or benadryl or benahist or bendylate or benekraft or benzhydramine or bromanate or calm-aid or derma-pax or dimedrol or dimine or diphen or diphenadryl or diphenhist or diphenhydramine or diphenmax or diphenyl or diphergan or diprazin or dormarex or dytan or dytuss or eldadryl or Fargan or Farganesse or genahist or hydramine or hyrexin or isopromethazine or Lergigan or medinex or nervine or nightcalm or nu-med or nytol or pardryl or paxidorm or pediacare or pentazine or phenadoz or phenazine or phendry or phenergan or phenerzine or phenoject or phensedyl or phenylbenzene or pipolphen or pro-med or proazamine or progan or promacot or promet or prometazin or promethegan or prorex or prothazin or Prothiazine or provigan or pyrethia or quenalin or remsed or Romergan or Receptozine or rumergan or siladryl or siladyl or silphen or sleep tab* or sleep-ettes or sleep-eze or sleepia or sleepinal or sominex or somnicaps or trux-adryl or tusstat or twilite or uni-hist or uni-tann or unisom sleepgels or unisom sleepmelts or valu-dryl or wehdryl or zipan).mp.  17. exp serotonin antagonist/  18. (serotonin adj2 antagonist*).tw,kw.  19. (dolasetron or anzemet).mp.  20. (granisetron or granisol or kytril or sancuso).mp.  21. (Ondansetron or zensana or zofran).mp.  22. (tropisetron or Navoban or Setrovel).mp.  23. exp Cholinergic Antagonists/ or anticholinergic agent*.tw.  24. (scopolamine or atrochin or boroscopol or buscapine or buscolysin or buscopan or butylscopolamine or butylscopolammonium bromide or epoxytropine tropate or hyocine hydrobromide or hyoscinbutylbromide or hyoscine or kwell or levo-duboisine or maldemar or scoburen or scopace or scopoderm or scopolaminebutylbromide or scopolaminum hydrobromicum or scopolan or transderm or transderm-scop or travacalm or vorigeno).mp.  25. exp antihistaminic agent/ or (histamine antagonist* or antihistamine* or anti-histamine*).mp.  26. (buclizine or cyclizine).tw.  27. (dimenhydrinate or antimo or aviomarin or biodramina or cinfamar or contramareo or dimen heumann or dimen lichtenstein or dimetabs or dinate or diphenhydramine theoclate or dramamine or dramin or Driminate or dramanate or dramoject or dymenate or gravol or Gravamin or marmine or nausicalm or reisegold or reisetabletten ratiopharm or reisetabletten stada or rodovan or rubiemen or superpep or travel-eze or travel-wise or triptone or uni-calm or Vertirosan or Viabom or vomex or vomacur or vomisin or wehamine).mp.  28. (Trimethobenzamide or barogan or benzacot or tebamide or ticon or tigan).mp.  29. (meclizine or agyrax or antivert or bonamine or bonikraft or bonine or chiclida or histametizyn or meclicot or meclozine or medivert or parachloramine or ruvertm or univert).mp.  30. exp Benzodiazepines/ or benzodiazepine*.mp.  31. (lorazepam or almazine or apolorazepam or ativan or donix or durazolam or idalprem or laubeel or lorazep or novolorazem or nuloraz or orfidal wyeth or sedicepan or sinestron or somagerol or tolid or temesta).mp.  32. exp corticosteroid/  33. corticosteroid*.tw,kw.  34. (dexamethasone or aacidexam or adexone or adrenocot or aeroseb or aknichthol dexa or alba-dex or alin or ambene or amplidermis or anemul mono or antimicotico or aquapred or auricularum or auxiloson or azona or baycadron or baycuten or cebedex or corson or cortastat or cortidex or cortidexason or cortisumman or corto-tavegil or dalalone or deca or decacort or decaderm or decadron or decalix or decasone or decaspray or dectancyl or deenar or dekasol or deronil or desamethasone or desameton or dexa-mamallet or dexa-rhinosan or dexa-scheroson or dexa-sine or dexacen or dexacort phosphate or dexacort* or dexafarma or dexafluorene or dexair or dexaject or dexalocal or dexamecortin or dexameth or dexamethasonedisodium phosphate or dexamethasonum or dexamonozon or dexapos or dexasol or dexasone or dexinoral or dexium or dexpak or dinormon or doxiproct or fluorodelta or fortecortin or gammacorten or hexadecadrol or hexadrol or lokalison or loverine or maxidex or medidex or metazone or methylfluorprednisolone or millicorten or mymethasone or ocasa or ocu-dex or oradexon or orgadrone or otomize or ozurdex or predni or primethasone or robadex or soludecadron or solurex or spersadex or trabit or visumetazone or voren).mp.  35. (methylprednisolone or a-methapred or adlone or ak-pred or ak-tate or aprednislon or articulose or asmacortone or balpred or blephamide liquifilm or bubbli-pred or caberdelta or capsoid or codelson or cortalone or corti-clyss or cortimed or cortisolone or cotolone or cryosolona or decaprednil or decortin or delta-cortef or delta-diona or delta-phoricol or deltacortilen or deltacortril or deltahydrocortisone or deltasolone or deltastab or deltidrosol or depmedalone or depo moderin or depo-medrol or depo-nisolone or depoject or depopred or dhasolone or di-adreson-f or diopred or dontisolon or duralone or duro cort or econopred or emmetipi or esametone or estilsona or firmacort or fisopred or flo-pred or frisolona or gupisone or hexacortone or hostacortin or hydeltra or hydeltrasol or hydrocortancyl or inf-oph or inflamase or inflanefran or isolone or key-pred or klismacort or kuhlprednon or lenisolone or lepi-cortinolo or locaseptil or longiprednil or med-jec or medicort or medlone or medrate or medrol or medrone or mega-star or meprdl or meprolone or metacortandralone or methacort or methylcotol or methylcotolone or methylone or methylpred or methylprednisolonum or meti derm or meticortelone or metilbetasone solubile or metipred or metrocort or metypresol or metysolon or millipred or ocu-pred or omnipred or ophtho-tate or opredsone or orapred or panafcortelone or pediapred or poly-pred liquifilm or polypred or precortalon aquosum or precortisyl or pred-clysma or pred-ject or pred-phosphate or predacorten or predair or predaject or predalone or predate or predcor or predeltilone or predenema or predfoam or predicort or predmix or prednabene or prednefrin or predni-coelin or predni or predni-helvacort or predni-m-tablinen or predni-pos or prednicortelone or prednihexal or prednilen or predniocil or prednisol or prednisolone or prednoral or predonine or predsol or prelone or pri-cortin or pri-methylate or pricortin or radilem or sano-drol or scherisolone-kristall or sieropresol or solpredone or solu moderin or solu-medrol or sterane or stintisone or summicort or urbason or urbasonsoluble or veripred or wyacort).mp.  36. exp cannabinoid/ or cannabinoid*.tw,kw.  37. (cannador or charas or ganja* or hashish or hemp or cannabis or marihuana or marijuana).tw,kw.  38. (marinol or dronabinol or tetrahydrocannabinol).mp.  39. or/8-38  40. (prevent* or mitigat* or minimi?* or avoid*).tw.  41. exp *prophylaxis/ or prophyla*.tw,kw.  42. 40 or 41  43. 7 and 39 and 42  44. animal/ not (animal/ and human/)  45. (veterinary or rabbit or rabbits or animal or animals or mouse or mice or rodent or rodents or rat or rats or murine or hamster* or pig or pigs or piglets or swine or porcine or horse* or equine or cow or cows or cattle or bovine or goat or goats or sheep or lambs or ovine or monkey or monkeys or trout or marmoset$1 or canine or dog or dogs or feline or cat or cats or zebrafish).ti.  46. 44 or 45  47. 43 not 46  48. limit 47 to "remove medline records" |
| **Cochrane Library**  via Wiley | #1 [mh "Deglutition Disorders"] or [mh Deglutition]  #2 [mh "Pneumonia, Aspiration"] or (aspiration NEXT pneumon*) or pneumonit* or (pulmonary NEXT aspiration*) or "foreign body aspiration"  #3 [mh "Respiratory Aspiration"]  #4 (aspiration NEXT pneumon*) or pneumonit* or (pulmonary NEXT aspiration*) or "foreign body aspiration" or (aspiration NEXT event*)  #5 ((airway* or respirat*) NEAR/3 aspirat*)  #6 ((swallow* or deglutit* or dysphag*) NEAR/3 (abnormal* or condition* or damage* or disturbance* or disorder* or difficult* or dysfunction* or impair* or injur*))  #7 ((throat or oesophag* or esophag* or pharyn* or oropharyn*) NEAR/3 (abnormal* or condition* or damage* or disturbance* or disorder* or difficult* or dysfunction* or impair* or injur*))  #8 {OR #1-#7}  #9 [mh Antiemetics] or antiemetic* or anti-emetic*  #10 [mh "Dopamine Antagonists"]  #11 dopamin* NEAR/2 antagonist*  #12 chlorpromazine or aminazine or chloractil or chlordelazine or contomin or dozine or fenactil or largactil or ormazine or propaphenin or thorazine  #13 domperidon* or domidon or evoxin or gastrocure or motilium or motillium or motinorm or costi or nauzelin  #14 metoclopramide or cerucal or clopra or degan or gastrese or gastrobid continus or gastroflux or gastromax or maxolon or maxeran or metaclopramide or metozolv or metramid or migravess or mygdalon or octamide or parmid or primperan or pylomid or reglan or reliveran or rimetin  #15 haloperidol or dozic or Aloperidin or Bioperidolo or Brotopon Duraperidol or fortunan or haldol or kentace or Einalon or Eukystol, or Halosten or Keselan or Linton or Peluces or Serenase or Sigaperidol or serenace  #16 prochlorperazine or buccastem or compazine or compro or emezine or procot or proziere or Phenotil or stemetil or Stemzine  #17 promethazine or Avomine or adgan or aler-dryl or aler-tab or aller-dryl topical or allergia or allermax or altaryl or anergan or antihist or antinaus or antituss or atosil or banaril or banophen or beldin or belix or ben tann or benadryl or benahist or bendylate or benekraft or benzhydramine or bromanate or calm-aid or derma-pax or dimedrol or dimine or diphen or diphenadryl or diphenhist or diphenhydramine or diphenmax or diphenyl or diphergan or diprazin or dormarex or dytan or dytuss or eldadryl or Fargan or Farganesse or genahist or hydramine or hyrexin or isopromethazine or Lergigan or medinex or nervine or nightcalm or nu-med or nytol or pardryl or paxidorm or pediacare or pentazine or phenadoz or phenazine or phendry or phenergan or phenerzine or phenoject or phensedyl or phenylbenzene or pipolphen or pro-med or proazamine or progan or promacot or promet or prometazin or promethegan or prorex or prothazin or Prothiazine or provigan or pyrethia or quenalin or remsed or Romergan or Receptozine or rumergan or siladryl or siladyl or silphen or sleep tab* or sleep-ettes or sleep-eze or sleepia or sleepinal or sominex or somnicaps or trux-adryl or tusstat or twilite or uni-hist or uni-tann or unisom sleepgels or unisom sleepmelts or valu-dryl or wehdryl or zipan  #18 [mh "Serotonin Antagonists"]  #19 (serotonin NEAR/2 antagonist*)  #20 dolasetron or anzemet  #21 granisetron or granisol or kytril or sancuso  #22 Ondansetron or zensana or zofran  #23 tropisetron or Navoban or Setrovel  #24 [mh "Cholinergic Antagonists"] or (anticholinergic NEXT agent*)  #25 scopolamine or atrochin or boroscopol or buscapine or buscolysin or buscopan or butylscopolamine or butylscopolammonium bromide or epoxytropine tropate or hyocine hydrobromide or hyoscinbutylbromide or hyoscine or kwell or levo-duboisine or maldemar or scoburen or scopace or scopoderm or scopolaminebutylbromide or scopolaminum hydrobromicum or scopolan or transderm or transderm-scop or travacalm or vorigeno  #26 [mh "Histamine Antagonists"] or (histamine NEXT antagonist*) or antihistamine* or anti-histamine*  #27 buclizine or cyclizine  #28 dimenhydrinate or antimo or aviomarin or biodramina or cinfamar or contramareo or dimen heumann or dimen lichtenstein or dimetabs or dinate or diphenhydramine theoclate or dramamine or dramin or Driminate or dramanate or dramoject or dymenate or gravol or Gravamin or marmine or nausicalm or reisegold or reisetabletten ratiopharm or reisetabletten stada or rodovan or rubiemen or superpep or travel-eze or travel-wise or triptone or uni-calm or Vertirosan or Viabom or vomex or vomacur or vomisin or wehamine  #29 Trimethobenzamide or barogan or benzacot or tebamide or ticon or tigan  #30 meclizine or agyrax or antivert or bonamine or bonikraft or bonine or chiclida or histametizyn or meclicot or meclozine or medivert or parachloramine or ruvertm or univert  #31 [mh Benzodiazepines] or benzodiazepine*  #32 lorazepam or almazine or apolorazepam or ativan or donix or durazolam or idalprem or laubeel or lorazep or novolorazem or nuloraz or orfidal wyeth or sedicepan or sinestron or somagerol or tolid or temesta  #33 [mh "Adrenal Cortex Hormones"]  #34 corticosteroid*  #35 dexamethasone or aacidexam or adexone or adrenocot or aeroseb or aknichthol dexa or alba-dex or alin or ambene or amplidermis or anemul mono or antimicotico or aquapred or auricularum or auxiloson or azona or baycadron or baycuten or cebedex or corson or cortastat or cortidex or cortidexason or cortisumman or corto-tavegil or dalalone or deca or decacort or decaderm or decadron or decalix or decasone or decaspray or dectancyl or deenar or dekasol or deronil or desamethasone or desameton or dexa-mamallet or dexa-rhinosan or dexa-scheroson or dexa-sine or dexacen or dexacort phosphate or dexacort* or dexafarma or dexafluorene or dexair or dexaject or dexalocal or dexamecortin or dexameth or dexamethasonedisodium phosphate or dexamethasonum or dexamonozon or dexapos or dexasol or dexasone or dexinoral or dexium or dexpak or dinormon or doxiproct or fluorodelta or fortecortin or gammacorten or hexadecadrol or hexadrol or lokalison or loverine or maxidex or medidex or metazone or methylfluorprednisolone or millicorten or mymethasone or ocasa or ocu-dex or oradexon or orgadrone or otomize or ozurdex or predni or primethasone or robadex or soludecadron or solurex or spersadex or trabit or visumetazone or voren  #36 methylprednisolone or a-methapred or adlone or ak-pred or ak-tate or aprednislon or articulose or asmacortone or balpred or blephamide liquifilm or bubbli-pred or caberdelta or capsoid or codelson or cortalone or corti-clyss or cortimed or cortisolone or cotolone or cryosolona or decaprednil or decortin or delta-cortef or delta-diona or delta-phoricol or deltacortilen or deltacortril or deltahydrocortisone or deltasolone or deltastab or deltidrosol or depmedalone or depo moderin or depo-medrol or depo-nisolone or depoject or depopred or dhasolone or di-adreson-f or diopred or dontisolon or duralone or duro cort or econopred or emmetipi or esametone or estilsona or firmacort or fisopred or flo-pred or frisolona or gupisone or hexacortone or hostacortin or hydeltra or hydeltrasol or hydrocortancyl or inf-oph or inflamase or inflanefran or isolone or key-pred or klismacort or kuhlprednon or lenisolone or lepi-cortinolo or locaseptil or longiprednil or med-jec or medicort or medlone or medrate or medrol or medrone or mega-star or meprdl or meprolone or metacortandralone or methacort or methylcotol or methylcotolone or methylone or methylpred or methylprednisolonum or meti derm or meticortelone or metilbetasone solubile or metipred or metrocort or metypresol or metysolon or millipred or ocu-pred or omnipred or ophtho-tate or opredsone or orapred or panafcortelone or pediapred or poly-pred liquifilm or polypred or precortalon aquosum or precortisyl or pred-clysma or pred-ject or pred-phosphate or predacorten or predair or predaject or predalone or predate or predcor or predeltilone or predenema or predfoam or predicort or predmix or prednabene or prednefrin or predni-coelin or predni or predni-helvacort or predni-m-tablinen or predni-pos or prednicortelone or prednihexal or prednilen or predniocil or prednisol or prednisolone or prednoral or predonine or predsol or prelone or pri-cortin or pri-methylate or pricortin or radilem or sano-drol or scherisolone-kristall or sieropresol or solpredone or solu moderin or solu-medrol or sterane or stintisone or summicort or urbason or urbasonsoluble or veripred or wyacort  #37 [mh cannabinoids] or cannabinoid*  #38 cannador or charas or ganja* or hashish or hemp or cannabis or marihuana or marijuana  #39 marinol or dronabinol or tetrahydrocannabinol  #40 {OR #9-#39}  #41 (prevent* or mitigat* or minimi?* or avoid*):ti,ab,kw  #42 prophyla*:ti,ab,kw  #43 #41 OR #42  #44 #8 AND #40 AND #43  #45 [mh animals] NOT ([mh animals] AND [mh humans])  #46 (veterinary or rabbit or rabbits or animal or animals or mouse or mice or rodent or rodents or rat or rats or murine or hamster* or pig or pigs or piglets or swine or porcine or horse* or equine or cow or cows or cattle or bovine or goat or goats or sheep or lambs or ovine or monkey or monkeys or trout or marmoset* or canine or dog or dogs or feline or cat or cats or zebrafish):ti  #47 #45 OR #46  #48 #44 NOT #47 |
| **CINAHL** | S1 (MH "Deglutition Disorders") OR (MH "Deglutition")  S2 (MH "Pneumonia, Aspiration")  S3 (MH "Aspiration")  S4 "aspiration pneumon*" or pneumonit* or "pulmonary aspiration*" or "foreign body aspiration" or "aspiration event*"  S5 (airway* or respirat*) N3 aspirat*  S6 ((swallow* or deglutit* or dysphag*) N3 (abnormal* or condition* or damage* or disturbance* or disorder* or difficult* or dysfunction* or impair* or injur*))  S7 ((throat or oesophag* or esophag* or pharyn* or oropharyn*) N3 (abnormal* or condition* or damage* or disturbance* or disorder* or difficult* or dysfunction* or impair* or injur*))  S8 S1 OR S2 OR S3 OR S4 OR S5 OR S6 OR S7  S9 (MH "Antiemetics+")  S10 antiemetic* or anti-emetic*  S11 (MH "Dopamine Antagonists+")  S12 dopamin* N2 antagonist*  S13 chlorpromazine or aminazine or chloractil or chlordelazine or contomin or dozine or fenactil or largactil or ormazine or propaphenin or thorazine  S14 domperidon* or domidon or evoxin or gastrocure or motilium or motillium or motinorm or costi or nauzelin  S15 metoclopramide or cerucal or clopra or degan or gastrese or gastrobid continus or gastroflux or gastromax or maxolon or maxeran or metaclopramide or metozolv or metramid or migravess or mygdalon or octamide or parmid or primperan or pylomid or reglan or reliveran or rimetin  S16 haloperidol or dozic or Aloperidin or Bioperidolo or Brotopon Duraperidol or fortunan or haldol or kentace or Einalon or Eukystol, or Halosten or Keselan or Linton or Peluces or Serenase or Sigaperidol or serenace  S17 prochlorperazine or buccastem or compazine or compro or emezine or procot or proziere or Phenotil or stemetil or Stemzine  S18 promethazine or Avomine or adgan or aler-dryl or aler-tab or aller-dryl topical or allergia or allermax or altaryl or anergan or antihist or antinaus or antituss or atosil or banaril or banophen or beldin or belix or ben tann or benadryl or benahist or bendylate or benekraft or benzhydramine or bromanate or calm-aid or derma-pax or dimedrol or dimine or diphen or diphenadryl or diphenhist or diphenhydramine or diphenmax or diphenyl or diphergan or diprazin or dormarex or dytan or dytuss or eldadryl or Fargan or Farganesse or genahist or hydramine or hyrexin or isopromethazine or Lergigan or medinex or nervine or nightcalm or nu-med or nytol or pardryl or paxidorm or pediacare or pentazine or phenadoz or phenazine or phendry or phenergan or phenerzine or phenoject or phensedyl or phenylbenzene or pipolphen or pro-med or proazamine or progan or promacot or promet or prometazin or promethegan or prorex or prothazin or Prothiazine or provigan or pyrethia or quenalin or remsed or Romergan or Receptozine or rumergan or siladryl or siladyl or silphen or sleep tab* or sleep-ettes or sleep-eze or sleepia or sleepinal or sominex or somnicaps or trux-adryl or tusstat or twilite or uni-hist or uni-tann or unisom sleepgels or unisom sleepmelts or valu-dryl or wehdryl or zipan  S19 (MH "Serotonin Antagonists+")  S20 serotonin N2 antagonist*  S21 dolasetron or anzemet  S22 granisetron or granisol or kytril or sancuso  S23 Ondansetron or zensana or zofran  S24 tropisetron or Navoban or Setrovel  S25 (MH "Cholinergic Antagonists+")  S26 "anticholinergic agent*"  S27 scopolamine or atrochin or boroscopol or buscapine or buscolysin or buscopan or butylscopolamine or butylscopolammonium bromide or epoxytropine tropate or hyocine hydrobromide or hyoscinbutylbromide or hyoscine or kwell or levo-duboisine or maldemar or scoburen or scopace or scopoderm or scopolaminebutylbromide or scopolaminum hydrobromicum or scopolan or transderm or transderm-scop or travacalm or vorigeno  S28 (MH "Histamine Antagonists+")  S29 "histamine antagonist*" or antihistamine* or anti-histamine*  S30 buclizine or cyclizine  S31 dimenhydrinate or antimo or aviomarin or biodramina or cinfamar or contramareo or "dimen heumann" or "dimen lichtenstein" or dimetabs or dinate or "diphenhydramine theoclate" or dramamine or dramin or Driminate or dramanate or dramoject or dymenate or gravol or Gravamin or marmine or nausicalm or reisegold or "reisetabletten ratiopharm" or "reisetabletten stada" or rodovan or rubiemen or superpep or travel-eze or travel-wise or triptone or uni-calm or Vertirosan or Viabom or vomex or vomacur or vomisin or wehamine  S32 Trimethobenzamide or barogan or benzacot or tebamide or ticon or tigan  S33 meclizine or agyrax or antivert or bonamine or bonikraft or bonine or chiclida or histametizyn or meclicot or meclozine or medivert or parachloramine or ruvertm or univert  S34 (MH "Antianxiety Agents, Benzodiazepine+")  S35 benzodiazepine*  S36 lorazepam or almazine or apolorazepam or ativan or donix or durazolam or idalprem or laubeel or lorazep or novolorazem or nuloraz or "orfidal wyeth" or sedicepan or sinestron or somagerol or tolid or temesta  S37 (MH "Adrenal Cortex Hormones+")  S38 corticosteroid*  S39 dexamethasone or aacidexam or adexone or adrenocot or aeroseb or "aknichthol dexa" or alba-dex or alin or ambene or amplidermis or "anemul mono" or antimicotico or aquapred or auricularum or auxiloson or azona or baycadron or baycuten or cebedex or corson or cortastat or cortidex or cortidexason or cortisumman or corto-tavegil or dalalone or deca or decacort or decaderm or decadron or decalix or decasone or decaspray or dectancyl or deenar or dekasol or deronil or desamethasone or desameton or dexa-mamallet or dexa-rhinosan or dexa-scheroson or dexa-sine or dexacen or dexacort phosphate or dexacort* or dexafarma or dexafluorene or dexair or dexaject or dexalocal or dexamecortin or dexameth or "dexamethasonedisodium phosphate" or dexamethasonum or dexamonozon or dexapos or dexasol or dexasone or dexinoral or dexium or dexpak or dinormon or doxiproct or fluorodelta or fortecortin or gammacorten or hexadecadrol or hexadrol or lokalison or loverine or maxidex or medidex or metazone or methylfluorprednisolone or millicorten or mymethasone or ocasa or ocu-dex or oradexon or orgadrone or otomize or ozurdex or predni or primethasone or robadex or soludecadron or solurex or spersadex or trabit or visumetazone or voren  S40 methylprednisolone or a-methapred or adlone or ak-pred or ak-tate or aprednislon or articulose or asmacortone or balpred or "blephamide liquifilm" or bubbli-pred or caberdelta or capsoid or codelson or cortalone or corti-clyss or cortimed or cortisolone or cotolone or cryosolona or decaprednil or decortin or delta-cortef or delta-diona or delta-phoricol or deltacortilen or deltacortril or deltahydrocortisone or deltasolone or deltastab or deltidrosol or depmedalone or "depo moderin" or depo-medrol or depo-nisolone or depoject or depopred or dhasolone or di-adreson-f or diopred or dontisolon or duralone or "duro cort" or econopred or emmetipi or esametone or estilsona or firmacort or fisopred or flo-pred or frisolona or gupisone or hexacortone or hostacortin or hydeltra or hydeltrasol or hydrocortancyl or inf-oph or inflamase or inflanefran or isolone or key-pred or klismacort or kuhlprednon or lenisolone or lepi-cortinolo or locaseptil or longiprednil or med-jec or medicort or medlone or medrate or medrol or medrone or mega-star or meprdl or meprolone or metacortandralone or methacort or methylcotol or methylcotolone or methylone or methylpred or methylprednisolonum or "meti derm" or meticortelone or "metilbetasone solubile" or metipred or metrocort or metypresol or metysolon or millipred or ocu-pred or omnipred or ophtho-tate or opredsone or orapred or panafcortelone or pediapred or "poly-pred liquifilm" or polypred or "precortalon aquosum" or precortisyl or pred-clysma or pred-ject or pred-phosphate or predacorten or predair or predaject or predalone or predate or predcor or predeltilone or predenema or predfoam or predicort or predmix or prednabene or prednefrin or predni-coelin or predni or predni-helvacort or predni-m-tablinen or predni-pos or prednicortelone or prednihexal or prednilen or predniocil or prednisol or prednisolone or prednoral or predonine or predsol or prelone or pri-cortin or pri-methylate or pricortin or radilem or sano-drol or scherisolone-kristall or sieropresol or solpredone or "solu moderin" or solu-medrol or sterane or stintisone or summicort or urbason or urbasonsoluble or veripred or wyacort  S41 (MH "Cannabinoids+")  S42 cannabinoid* or cannador or charas or ganja* or hashish or hemp or cannabis or marihuana or marijuana  S43 marinol or dronabinol or tetrahydrocannabinol  S44 S9 OR S10 OR S11 OR S12 OR S13 OR S14 OR S15 OR S16 OR S17 OR S18 OR S19 OR S20 OR S21 OR S22 OR S23 OR S24 OR S25 OR S26 OR S27 OR S28 OR S29 OR S30 OR S31 OR S32 OR S33 OR S34 OR S35 OR S36 OR S37 OR S38 OR S39 OR S40 OR S41 OR S42 OR S43  S45 TI ( prevent* or mitigat* or minimi* or avoid* or prophyla* ) OR AB ( prevent* or mitigat* or minimi* or avoid* or prophyla* )  S46 (MH "Animals+")  S47 (MH "Human")  S48 S46 NOT ( (S46 AND S47) )  S49 TI (veterinary or rabbit or rabbits or animal or animals or mouse or mice or rodent or rodents or rat or rats or murine or hamster* or pig or pigs or piglets or swine or porcine or horse* or equine or cow or cows or cattle or bovine or goat or goats or sheep or lambs or ovine or monkey or monkeys or trout or marmoset* or canine or dog or dogs or feline or cat or cats or zebrafish)  S50 S48 OR S49  S51 S8 AND S44 AND S45  S52 S51 NOT S50 |

Table 3s: Risk of bias assessment for randomized control studies

| Study | Domain 1: Randomization Process | Domain 2: Deviations from the intended interventions | Domain 3: Missing outcome data | Domain 4: Measurement of outcome | Domain 5: Selection of the reported result | Overall ROB |
| --- | --- | --- | --- | --- | --- | --- |
| Allami  2022 | LR | LR | LR | LR | SC | LR |
| Warusevitane 2015 | LR | LR | SC | LR | LR | LR |
| Yavagal 2000 | LR | LR | LR | LR | LR | LR |
| LR: Low risk; SC: Some concerns; HR; High risk | | | | | | |
